# Clinical evaluation of a SARS-CoV-2 RT-PCR assay on a fully automated system for rapid on-demand testing in the hospital setting

**DOI:** 10.1101/2020.04.07.20056234

**Authors:** Dominik Nörz, Nicole Fischer, Alexander Schultze, Stefan Kluge, Ulrich Mayer-Runge, Martin Aepfelbacher, Susanne Pfefferle, Marc Lütgehetmann

## Abstract

1

**Background:** The ongoing SARS-CoV-2 pandemic presents a unique challenge for diagnostic laboratories around the world. Automation of workflows in molecular diagnostics are instrumental for coping with the large number of tests ordered by clinicians, as well as providing fast-tracked rapid testing for highly urgent cases. In this study we evaluated a SARS-CoV-2 LDT for the NeuMoDx 96 system, a fully automated device performing extraction and real-time PCR.

**Methods:** A publicly available SARS-CoV-2 RT-PCR assay was adapted for the automated system. Analytical performance was evaluated using in-vitro transcribed RNA and clinical performance was compared to the cobas 6800-based reference assay within the lab.

**Results:** The NeuMoDx-sarbeco-LDT displayed good analytical performance with an LoD of 95.55 cp/ml and no false positives during evaluation of cross-reactivity. A total of 176 patient samples were tested with both the Sarbeco-LDT and the reference assay. Positive and negative agreement were 100% and 99.2% respectively. Invalid-rate was 6.3%.

**Conclusion:** The NeuMoDx-sarbeco-LDT showed analytical and clinical performance comparable to the cobas6800-based reference assay. Due to its random-access workflow concept and rapid time-to-result of about 80 minutes, the device is very well suited for providing fast-tracked SARS-CoV-2 diagnostics for urgent clinical samples in the hospital setting.

**Highlights:**

- A publicly available SARS-CoV-2 RT-PCR assay was adapted and evaluated on the open mode of the NeuMoDx 96 system (Qiagen)
- The assay showed comparable analytical and clinical performance to the reference assay
- Fast turn-around times (80 minutes) and random-access workflow of the system makes the assay well suited for urgent clinical samples.

## 2 Introduction

In early January 2020, SARS-CoV-2 was first identified as the likely causative agent of a cluster of cases of viral pneumonia in the city of Wuhan, China (1). The novel virus is situated in the ‘sarbecovirus’ subgenus along with its genetically distinct relative, the original SARS-coronavirus (2). SARS-CoV-2 saw rapid spread worldwide eventually prompting the WHO to declare a ‘global health emergency’ by the end of January (3).

Outbreak scenarios present a unique challenge for diagnostic laboratories. Particularly in the case of respiratory viruses such as SARS-CoV-2, clinical symptoms can be largely indistinguishable from other common respiratory pathogens such as e.g. Influenza (4) and polymerase chain reaction (PCR) assays are necessary to confirm or rule out the novel virus (5). A variety of suitable assays were made available early on during the outbreak, notably by Corman et al. (6) and the CDC, which were swiftly adopted by many labs in Europe and around the world. However, their overall testing capacity remained limited (7). We and others have previously demonstrated how automation in molecular diagnostics enables easy scaling of testing capacity by substantially cutting back hands-on time for PCR-assays (8, 9).

For the assay presented in this study, we used a fully automated random-access platform for molecular diagnostics, handling everything from extraction, amplification, signal detection to reporting of results (10). For RNA targets, the time-to-result is approximately 80 minutes, given optimal conditions. The availability of an open mode allows for the rapid implementation of lab developed tests (LDT). The aim of this study was to adapt and evaluate a previously published SARS-CoV-2 PCR assay (by Corman et al. (6)) for the NeuModx 96/288 system.

## 3. Sarbeco-LDT assay setup

Primers (fwd: 5′-ACAGGTACGTTAATAGTTAATAGCmGT-3′, rev 5′-ATATTGCAGCAGTACGCACAmCA-3′) and probe (5′-Fam-ACACTAGCC/ZEN/ATCCTTACTGCGCTTCG-Iowa Black FQ-3’) used for Sarbeco-LDT were custom made and purchased from IDT DNA Technologies (Coralville, USA). Both primers were modified with 2’-O-methyl bases in their penultimate base to prevent formation of primer dimers (mG or mC). A double-quenched probe was used in order to reduce background fluorescence.

In accordance with instructions issued by the manufacturer, a 6x Primer/Probe mix was prepared and 5μL of the mix were loaded into the LDT-Strip well by well for each reaction (e.g. 400nM primers, 75 nM probe per reaction). For a complete run protocol see the test-summary displayed in table 1.

**Table 1:**
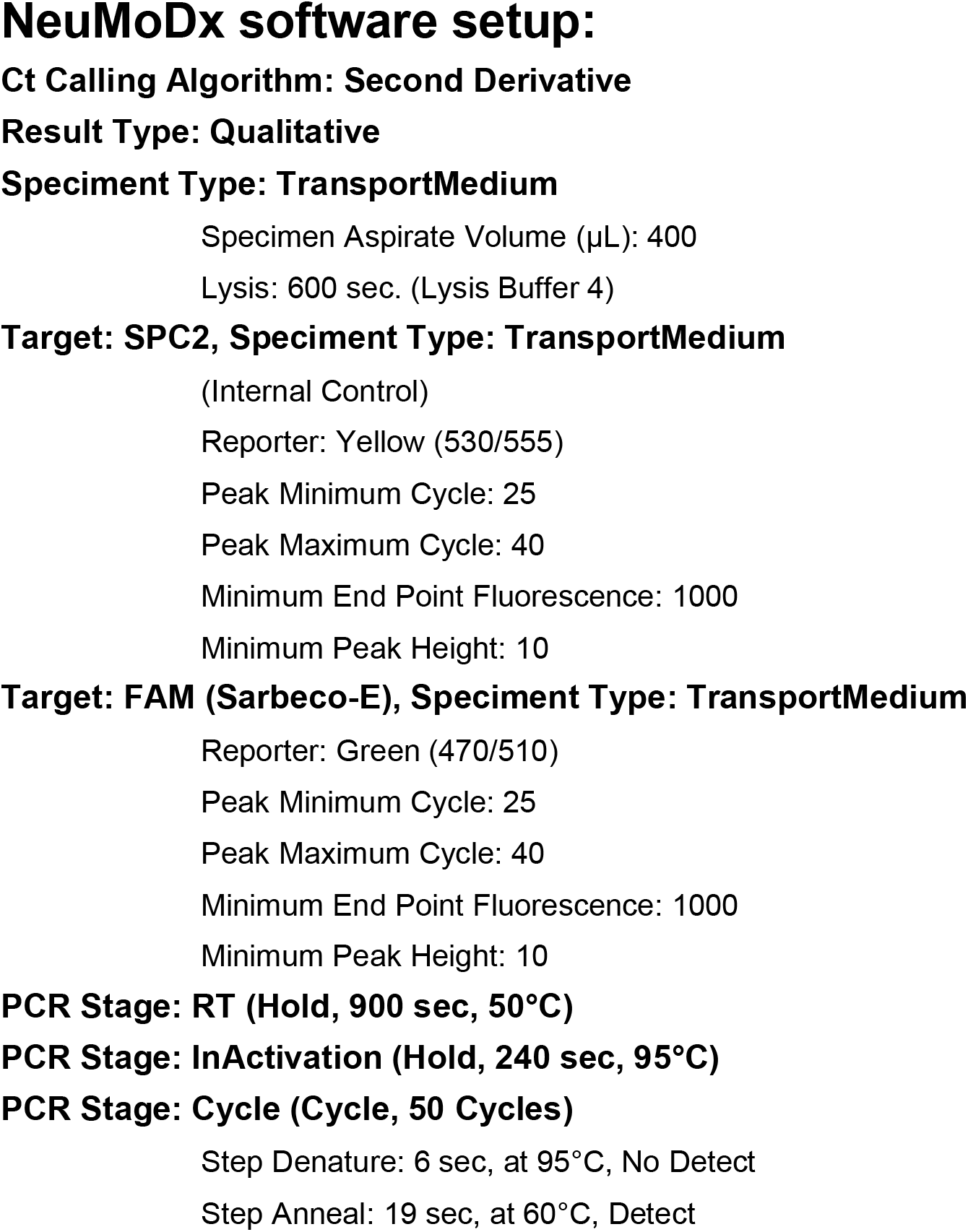
NeuMoDx-Software run-protocol summary displaying settings and PCR protocol.

Clinical specimens used for this study were oropharyngeal and nasopharyngeal swabs (E-Swab collection kits, Copan, Italy). Prior to analysis, 1ml Roche cobas PCR medium (≤ 40% guanidine hydrochloride in Tris-HCL buffer) was added to the sample in order to inactivate potential pathogens within and facilitate further handling. Samples were then briefly vortexed before being loaded into the instrument.

## 4 Assessment of analytical performance

For analytical evaluation, in-vitro transcribed RNA (IVT-RNA) of the viral E-gene was generated as described previously (6) using the following primers: 5′-TACTAATACGACTCACTATAGATACAGGTACGTTAATAGTTAATAGCGT-3′ and 5′-ttttttttgtatacATATTGCAGCAGTACGCACACA-3′. IVT-RNA was adjusted for copy-numbers to a predefined RNA standard obtained from “European virus archive” (EVA), (https://www.european-virus-archive.com).

A total of 8 replicates of 4 different concentrations (400, 100, 40 and 10 copies/ml) and negative control were used to determine LoD by probit-analysis (MedCalc, MedCalc Software Ltd). Limit of detection was determined as 95.55 cp/ml at 95% probability of detection (CI 63.56 cp/ml – 241.46 cp/ml). (Figure 1)

**Figure 1:**
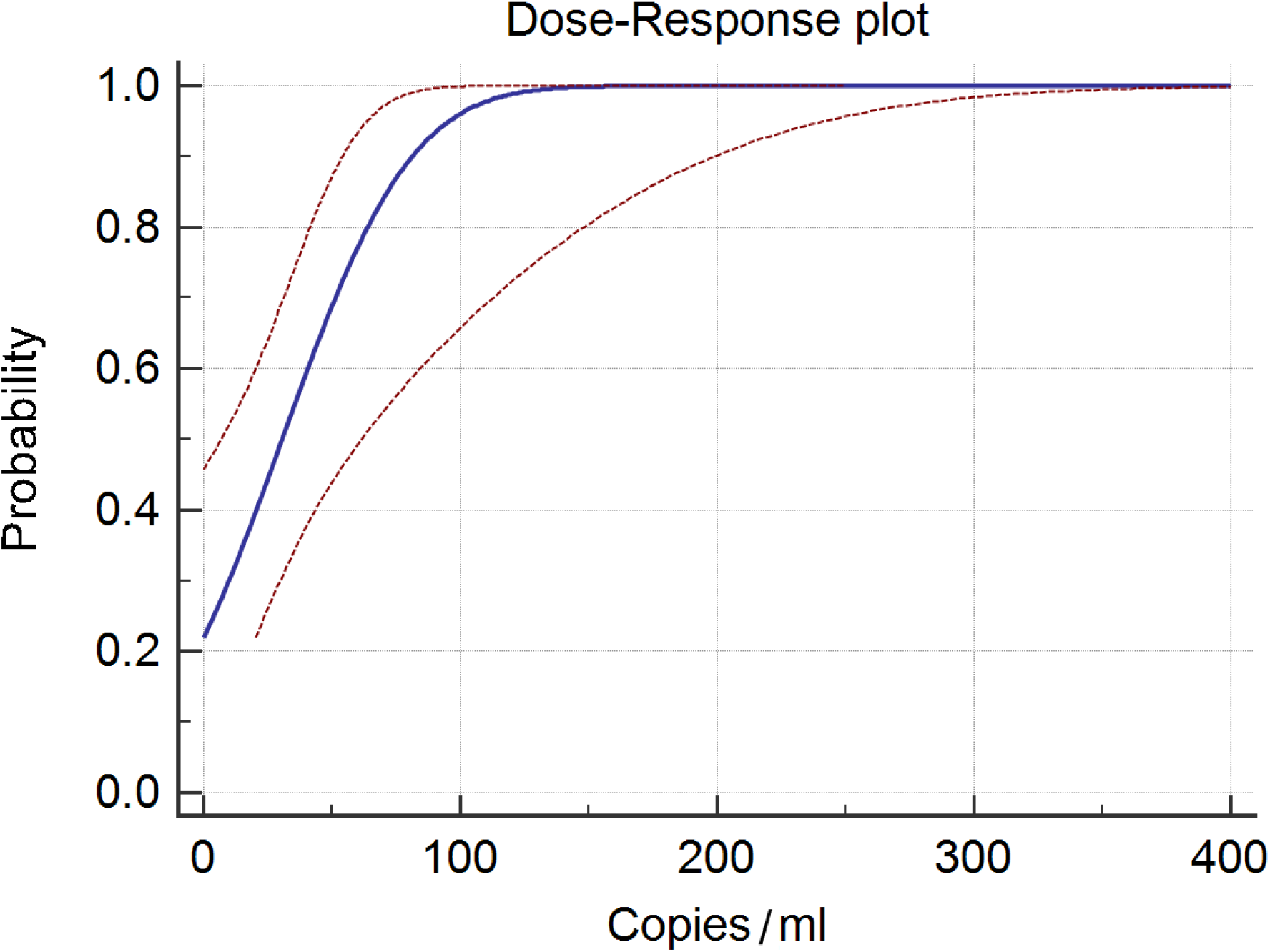
Probit-curve of LoD assessment (Blue curve, Probability of detection; Red curves, 95% confidence interval)

Inter-run and intra-run variability were evaluated using spiked swab samples containing IVT-RNA at approximately 5x and 10x LoD, running 5 repeats each on two different days. Median Ct values were 27.045 (+/- 0.695 ct) and 27.640 (+/- 1.14 ct) for 10x LoD and 5x LoD respectively.

In order to rule out potential cross-reactivity with other organisms present in respiratory swabs, a set of predetermined clinical samples containing a variety of respiratory pathogens and external quality assessment panel samples were selected and subjected to the Sarbeco-LDT. There were no false positive results, see table 2.

**Table 2:**
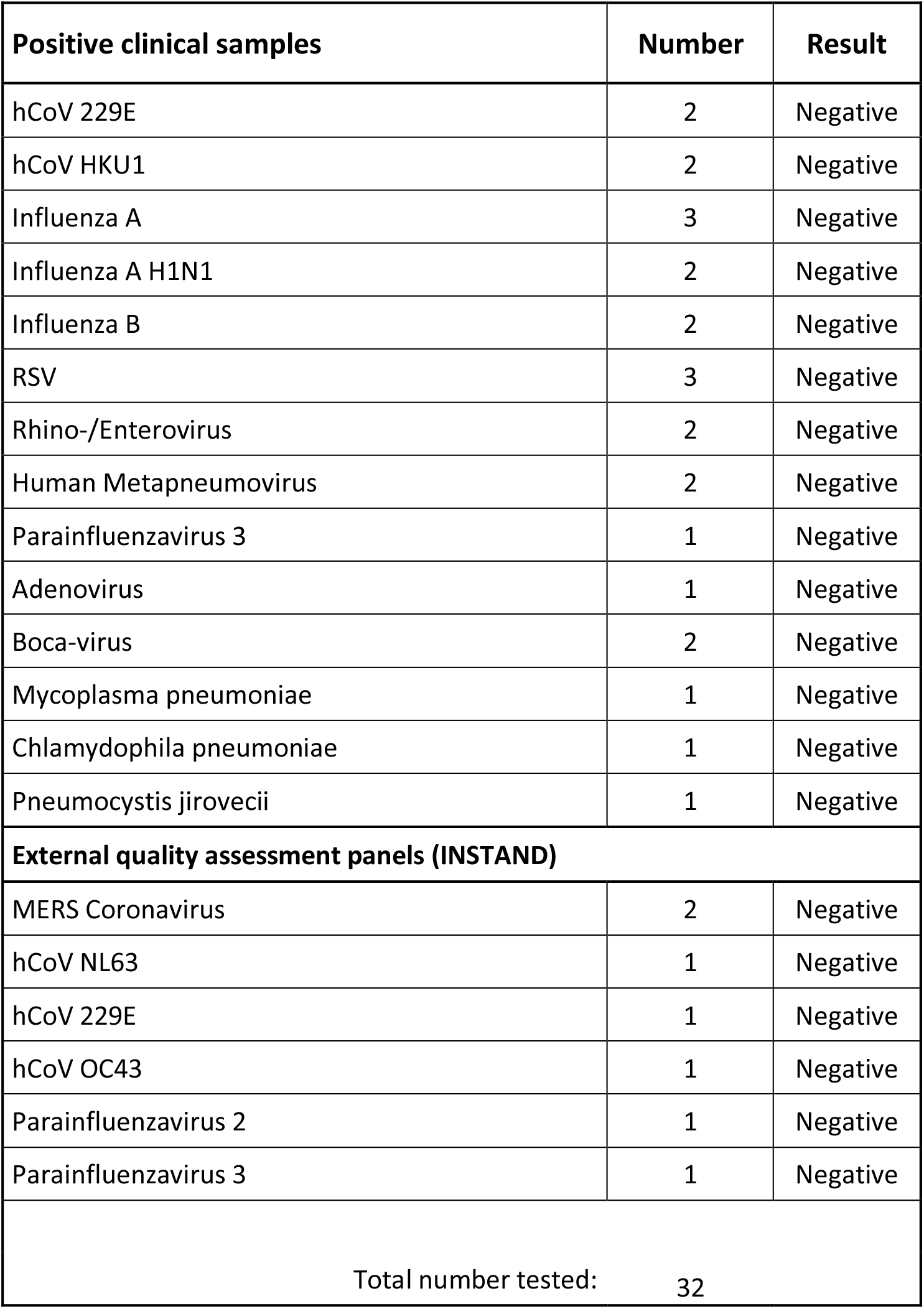
Clinical samples and external quality control samples (provided by INSTAND e.V., Düsseldorf, Germany) were tested for potential cross-reactivity with the Sarbeco-LDT.

## 5 Comparing clinical performance

Clinical performance of the assay was analyzed by comparing the Sarbeco-LDT to the reference method within the lab, the cobas6800-based “SARS-CoV-2 UCT” assay (11). A total of 176 clinical samples (collected during the time between 17/03/20 and 30/03/20) were prepared according to the above-mentioned protocol, split into aliquots and tested in parallel on both systems. Samples that did not yield valid results on the NeuMoDx system are reported as “Invalid”. The inhibition rate was 6.3% (11/176 samples, all of which were tested negative in the reference assay). Positive agreement was 100% (35/35, amplification curves see figure 2) and negative agreement was 99.2% (129/130). A single discrepant sample occurred, returning positive on the NeuMoDx system (late ct, close to LoD) and negative on the cobas6800. Root cause investigation revealed that this patient had previously been diagnosed with COVID-19 elsewhere.

**Figure 2:**
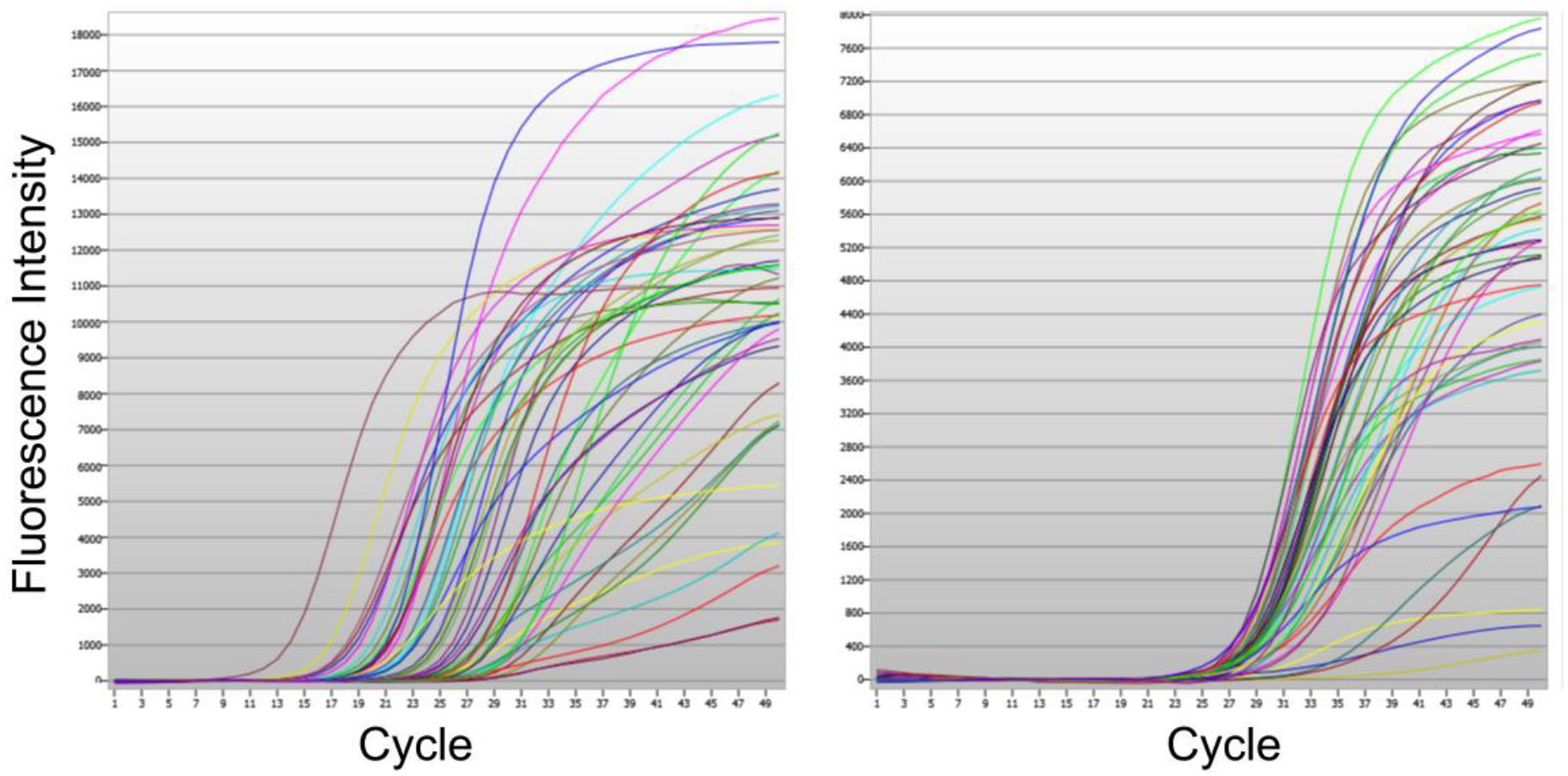
Amplification curves of all positive clinical samples as displayed by the NeuMoDx Software. Left: FAM-probe (E-gene), normalized; Right: VIC/JOE-probe (IC), normalized.

## 6 Discussion

When comparing the current SARS-CoV-2 outbreak to the SARS pandemic in 2003/04, it is immediately apparent how much faster emerging pathogens can be identified and characterized in the modern day (2, 12). TaqMan based RT-PCR-assays for the novel virus were available online mere days after the initial sequence of SARS-CoV-2 had been published (6). However, while these assays can be implemented relatively swiftly by local diagnostic laboratories, their reliance on manual PCR setups sets narrow limits to overall capacity. A study by Reusken et al. reported readiness to test for the novel Coronavirus by the end of January 2020 in almost all countries of the European union, but with a capacity of 250 tests per week or less for the vast majority of them (7). Similar issues were reported early on in China, where testing could not be performed for all suspected cases due to limitations in capacity (13).

In a recent study we demonstrated that a previously published TaqMan based SARS-CoV-2 RT-PCR assay, endorsed by ECDC and WHO, can be adapted to run on an automated batch-based high-throughput system, the cobas6800 (11). Utilizing this assay, more than 10,000 samples were tested for SARS-CoV-2 during the month of March 2020 while maintaining all other routine diagnostics in our laboratory, proving the potency of rapid automation to cope with massive surges in demand. However, taking into consideration sample registration, pretreatment, preparation of batches, and generating reports, it usually takes more than 5 hours before results can be made available to clinicians (14). Consequently, alternative workflows are required to enable fast-tracking of high-priority samples.

The NeuMoDx 96 system is a fully automated RT-PCR platform, performing extraction, amplification and signal detection without requiring any human interaction. it provides random-access capabilities and turn-around times of 80 minutes for RNA targets. In this study we have adapted the SARS-CoV-2 RT-PCR assay by Corman et al. (6) for use on the NeuMoDx 96 automated system. Analytical and clinical performance was comparable to the cobas6800-based reference assay (11), showing an LoD of approximately 100 copies/ml and positive and negative agreement of 100% and 99.2% respectively. The relatively high inhibition rate of 6.3% suggests that sample preparation procedures can be optimized further. The NeuMoDx-based Sarbeco-LDT represents a valuable complement to routine SARS-CoV-2 testing by offering the ability to run individual high-priority samples at any time and reporting results within two hours if necessary.

## 7 Conclusion

In this study we have adapted a publicly available SARS-CoV-2 screening assay for use on the open channel of the NeuMoDx 96/288 system (Qiagen). The assay demonstrates comparable analytical and clinical performance to established LDTs currently in use for SARS-CoV-2 diagnostics. Due to its random-access capabilities and short turn-around times (80 minutes), the system is well suited for automating medium-throughput routine SARS-CoV-2 testing, or as an addition to high-throughput systems to allow fast-tracking for highly urgent clinical samples.

## Data Availability

Data is available on request.

